# Does trust in government improve Covid-19’s crisis management?

**DOI:** 10.1101/2021.07.10.21260297

**Authors:** Ablam Estel Apeti

**Affiliations:** Université Clermont Auvergne, CNRS, IRD, CERDI, F-63000 Clermont-Ferrand, France

**Keywords:** Covid-19, containment measures, trust in government, recovery plans

## Abstract

Countries have adopted several measures to control the spread of Covid-19. However, substantial differences remain in terms of performance in controlling the virus, potentially due to heterogeneity in citizen engagement with government measures. Drawing on this observation, this paper seeks to analyze the effect of pre-crisis ties, particularly trust in government, on crisis management, proxied by the number of Covid-19 cases and deaths per million population. We examine this question based on a sample of 41 countries for which data are available. Results reveal that a high level of trust in government predicts better crisis management in terms of relatively low levels of cases and deaths. These results, which successfully pass a series of robustness tests, may vary according to level of contamination and increase with time.

**JEL Classification:** E71, H12, I12, I18, I38, Z18

## 1 Introduction

In response to Covid-19, most countries have implemented non-pharmaceutical interventions ranging from physical distancing to non-essential (internal) movement restrictions, i.e., lockdown, ^1^ and more recently, vaccination campaigns, to control the virus and save lives. Although the effect of these policies remains relatively positive on pandemic management (Acemoglu et al., 2020; Caselli et al., 2020; Cowling et al., 2020; Demirguc-Kunt et al., 2020), the levels of cases and deaths — proxy of crisis management — due to Covid-19 vary across countries or regions. ^2^ This heterogeneity in the crisis management indicates that compliance to policies put in place by governments may depend on some countries’ characteristics, especially citizens’ commitment. In particular, the ability of people to comply with government policies, and consequently the government’s ability to control the pandemic, may depend, like any other public policy response, on citizens’ trust in their government. ^3^

Although the literature on the Covid-19 pandemic remains sizeable, little is said about the role of trust in government ^4^ on the management of the crisis in terms of the number of cases and deaths. It is important to note that one paper that addresses a similar question is Gelfand et al. (2021). However, a major difference exists. Indeed, these authors analyze the effect of cultural tightness-looseness on the number of cases and deaths, while our goal is to focus on citizens’ trust in their government. ^5^ In most existing papers, trust is presented as a determinant of compliance to government health policies and hardly as a determinant of mortality, or more importantly, of the virus’ spread, i.e., contaminations or number of cases. This paper will fill this gap by analyzing the effect of pre-Covid trust in government on crisis management based on the number of cases and deaths per million population. Our results based on Ordinary Least Squares (OLS) on 41 countries show that high pre-crisis trust predicts better crisis management through lower numbers of cases and deaths per million population. This finding remains robust to alternative crisis management measures such as numbers of new cases and deaths per million population, number of hospitalized patients per million population, number of Intensive Care Unit (ICU) patients per million population, excess mortality during the pandemic, and Covid Performance Index. In addition, robustness tests performed by altering the sample, using alternative definitions of trust, and adding additional controls failed to alter our conclusions. The heterogeneity tests conducted later reveal that the trust effect may depend on the level of contamination, with a larger magnitude above 100 cases per million population, and increases over time i.e., between the first (2020) and second (2021) years of the pandemic.

The rest of the paper is organized as follows. Section 2 presents the arguments linking trust and Covid-19 management. Section 3 discusses the data and some descriptive statistics. Section 4 presents the methodology. Sections 5 and 6 discuss the results and robustness tests respectively. Section 7 highlights some conditions under which our baseline findings may vary, and Section 8 concludes.

## 2 The argument

Three main arguments support the idea that the pre-Covid trust can influence pandemic management.

First, trust can support the creation of broad (pre-crisis) policy space for government intervention related to Covid-19. ^6^ Various studies, including Schaltegger and Torgler (2005) and OECD (2013) show a positive association between trust and fiscal discipline in terms of lower public debt accumulation. Illustrative examples are the Nordic countries, in particular Denmark and Sweden, which exhibit high levels of trust and fiscal discipline (low debt) compared to most developed countries. ^7^ This pre-pandemic fiscal discipline may ease the creation of significant fiscal space for stimulating the economy, providing transfers and social assistance in order to mitigate crisis collateral damage ^8^ and amplifying factors such as unemployment, mental health problems, suicides and many others (Chiappini et al., 2020; Holman et al., 2020; Kola, 2020; Panchal et al., 2020; Silverio-Murillo et al., 2021). ^9^

Second, trust can help manage the pandemic by ensuring public compliance with government-announced health policies. As previously reported, in response to the rapid spread and growing death toll, and due to limited available and/or “widely” approved therapies, most governments have implemented non-pharmaceutical measures such as physical distancing, travel restrictions/bans, masks, stay-at-home orders, or regular hand washing. However, compliance with these strict measures that change the way millions of people live is not always deliberate. For example, in some parts of the world, these measures are perceived as a clear desire of (national) governments to control and restrict individual liberties, leading to protests, boycotts, and, to some extent, civil unrest. Existing studies show that compliance to these measures depends on people’ trust in their government (Yaqub et al., 2014; Blair et al., 2017; Bargain and Aminjonov, 2020). Thus, trust appears to play a key role in successful containment programs and potentially in the willingness of individuals to believe in science and adhere to new measures, mainly pharmaceutical such as vaccination, to tackle the virus, as documented in several studies (Woskie and Fallah, 2019; Deb et al., 2020; Hosny, 2021; Pagliaro et al., 2021).

Finally, trust can undermine sound management of the pandemic, by, for example, promoting less skilled governments. Indeed, excessive trust can lead citizens to naively believe that government is effectively managing the pandemic when it is not (Devine et al., 2020), possibly creating excessive levels of infections and deaths. On the other hand, low trust of citizens in politicians may favor populist parties (Keefer et al., 2021), which to date has shown less efficiency in managing the pandemic (example of Brazil).

In summary, putting these three arguments together, we can assume that the effect of trust on crisis management can be transmitted through three channels, namely the creation of relevant pre-Covid economic policy space for quick reaction in response to the crisis, the commitment of citizens to comply with government health policies or virus control measures, and the cover-up of government’s real capacity to effectively manage the health crisis and/or the implications of populist parties’ expansion. The contradiction revealed by these three channels shows that the effect of trust on the pandemic’s management is ambiguous, making the question more empirical than theoretical.

## 3 Data and first impressions

In this section, we describe the key variables in our paper and underscore some statistical evidence that characterizes the relationship between trust and Covid-19 crisis management.

### 3.1 Data

Our analysis is based on cross-sectional data of 41 countries with a geographical scope covering the five continents and including the following variables:

#### Trust in government

Trust in government refers to the share of people who report having confidence in the national government. The data shown reflect the share of respondents answering “yes” (the other response categories being “no”, and “don’t know”) to the survey question: “In this country, do you have confidence in … national government?”. The sample is ex-ante designed to be nationally representative of the population aged 15 and over. This indicator is measured as a percentage of all survey respondents. For our analysis, we use pre-Covid trust chiefly to remove any potential effects of the crisis management and thus assess the role of differences in structural characteristics such as norms, values, and especially trust on the pandemic management pathway. ^10^ For our primary measure, we consider average trust over 2018-2019. To test the robustness of our results, we used alternative trust measures by taking an average over the period 2006-2019, 2006-2019 without the Global Financial Crisis (GFC) period, using only 2019 observations. The data are from OECD (2021). In addition, we used the European Social Survey (ESS) database, which allows us to define three alternative trust measures: trust in politicians, citizens’ satisfaction with the work of the national government, and citizens’ satisfaction with the way democracy works in their country.

#### Covid-19 crisis management

We use two main measures for crisis management, namely the (total) number of cases and deaths per million population (virus prevalence). For robustness concerns, we use alternative measures of crisis management. First, we use pandemic incidence instead of prevalence captured by the number of new cases and deaths per million population. Second, we use the number of hospitalizations and Intensive Care Unit (ICU) patients admissions due to Covid-19. Third, we select excess mortality during the Covid-19 crisis, in particular P-score, which captures how the number of weekly or monthly deaths in 2020-2021 differs as a percentage from the average number of deaths during the same period over the years 2015-2019. Put another way, excess mortality is an epidemiology and public health term that refers to the number of deaths from all causes occurring during a crisis that exceed what would have been expected under “normal” conditions. In this case, we are interested in comparing the number of deaths that occurred during the Covid-19 pandemic to the number of deaths we would have expected had the pandemic not occurred — a crucial quantity that cannot be known but can be estimated in several ways. Excess mortality mitigates the number of deaths restriction by taking into account the total impact of the pandemic on deaths instead of solely Covid-19 confirmed deaths. Specifically, this variable takes into account not only confirmed deaths, but also Covid-19 deaths that were not properly diagnosed and reported, as well as deaths due to other causes that are attributable to the general conditions of the crisis. ^11^ The data come from Hannah Ritchie and Roser (2020). Finally, we pick Lowy Institute’s Covid Performance Index (CPI) as an alternative proxy of crisis management. This measure provides a ranked comparison of the performance of countries in managing the Covid-19 pandemic in the 43 weeks following their hundredth confirmed case of the virus, using data available to March 13, 2021 and is computed based on the following variables: confirmed cases, confirmed deaths, confirmed cases per million people, confirmed deaths per million people, confirmed cases as a proportion of tests, and tests per thousand people.

#### Macroeconomic variables

Macroeconomic variables from World Development Indicators (WDI) include data on real GDP per capita growth and trade openness.

#### Demographic and fiscal variables

Data from the WDI and the IMF’s World Economic Outlook (WEO) databases include population density and public expenditure over GDP.

#### Health preparedness

Data on hospital beds per thousand inhabitants are obtained from Hannah Ritchie and Roser (2020). Descriptions and sources for every variable used in this paper, as well as the sample composition, are provided in the Appendix.

### 3.2 First impressions

To get an idea on the relationship between trust ^12^ and Covid-19 crisis management, we start with some statistical regularity. Figure 1 shows a correlation between trust and the number of Covid-19 cases on the one hand, and between trust and number of deaths on the other hand. Irrespective of the measure of crisis management used, a negative association with trust is observed. In other words, a high pre-crisis level of trust in government is associated with better management of the Covid-19 crisis materialized by a relatively low number of cases and deaths. Table 1 extends this observation by computing the number of cases and deaths according to the level of trust. ^13^ The results reveal that countries in which citizens have high trust in government are dealing well with the crisis. Specifically, high trust countries exhibit contaminations (deaths) per million population of 18211.73 (333.90) versus 22764.65 (609.55) for low trust countries. Compared to low trust countries, these results indicate that high trust countries experience 20% and 45.22% reduction in contaminations and deaths over January 1, 2020 to May 26, 2021.

**Table 1.**
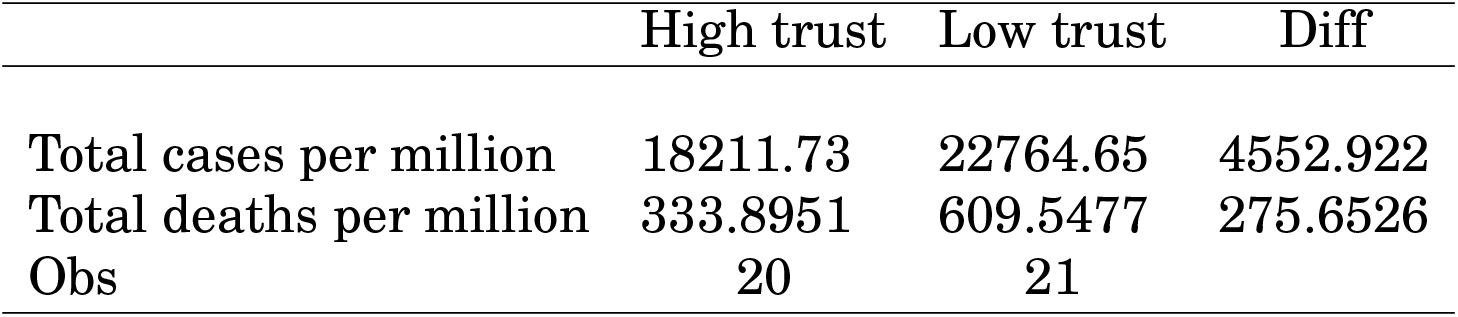
Covid-19 crisis management by trust level

| Table 1 – Covid-19 crisis management by trust level |  |  |  |
| --- | --- | --- | --- |
|  | High trust | Low trust | Diff |
| Total cases per million | 18211.73 | 22764.65 | 4552.922 |
| Total deaths per million | 333.8951 | 609.5477 | 275.6526 |
| Obs | 20 | 21 |  |

**Figure 1.**
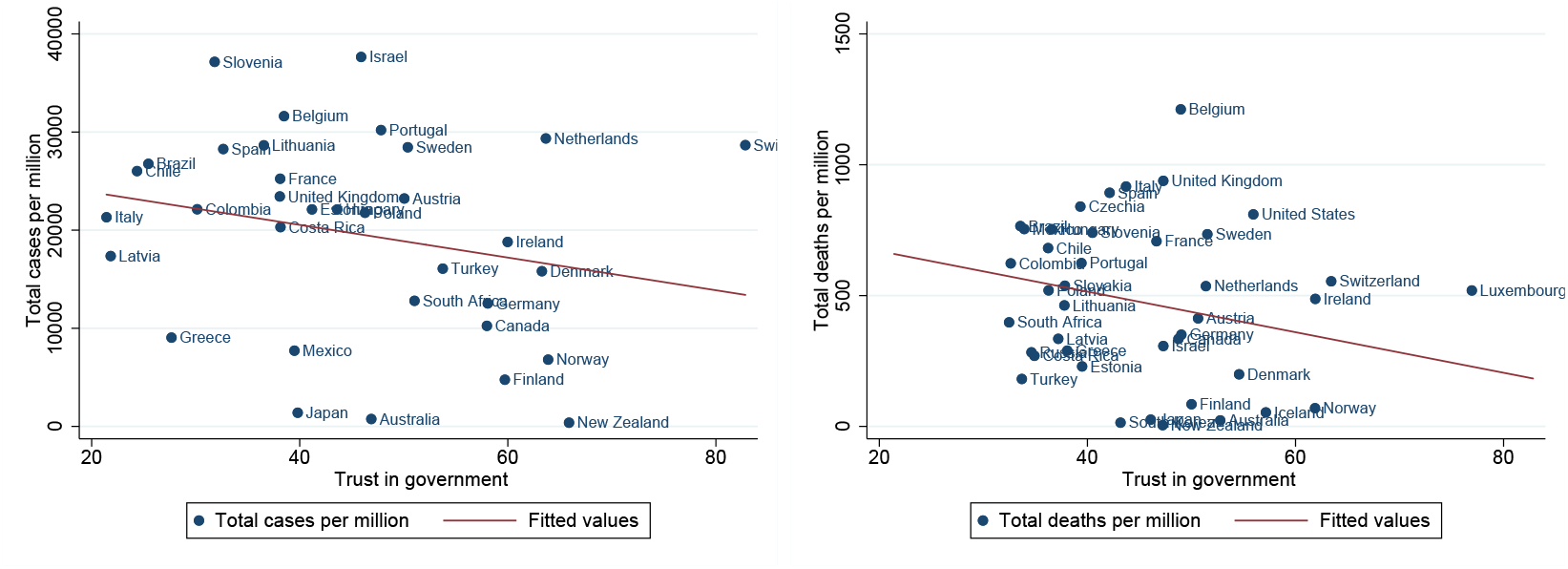
*Corelation between trust and Covid-19 crisis management* Source: Authors’ calculations

Keeping these descriptive relationships in mind, we develop a more formal analysis in the following section to capture the causal effect of trust, i.e., the impact on the Covid-19 crisis management signaled by the number of cases and deaths per million population.

## 4 Methodology

In this section, we first present the model used to test our question and then we develop our identification strategy.

### 4.1 Model

We estimate the effect of trust on the Covid-19 crisis management using a cross-section model :

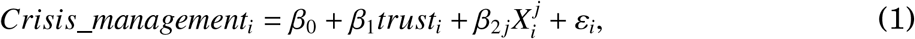

with *Crisis*_*management*_*i*_ the Covid-19 crisis management of country *i, trust*_*i*_ the (pre-Covid) trust in government of country *i*, 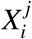 the vector of *j* control variables, and *ε*_*i*_ the error term.

The choice of control variables, including population density, public expenditure, hospital beds per thousand, trade openness, and real GDP per capita growth, is based on their potential effect on trust but also on the pandemic management. For example, high population density may lead to rapid circulation of the virus due to strong social or physical interaction, making crisis management more difficult. This is the typical case of large cities which generally exhibit large size of the crisis. On the other hand, good pre-crisis public expenditure and hospital bed availability can improve countries’ preparedness to manage the crisis. Meanwhile, good pre-pandemic economic conditions as tracked by real GDP per capita growth can shape pre-existing trust between politicians or policy-makers and citizens. The effect of trade openness may seem ambiguous. Indeed, trade openness can improve countries’ economic situation and thus create a strong trust relationship. On the other hand, high openness can result in strong dependence on foreign countries for some essential goods needed for managing the pandemic, such as masks, hydroalcoholic gels, or drugs (Barlow et al., 2021). This is the case of some countries that suffered, especially at the beginning of the pandemic, to have the necessary tools for the protection of their population and the limitation of the spread of the virus. The parameter of interest is *β*_1_: to confirm our hypothesis, *β*_1_ should be statistically significant and indicate a favorable effect of higher pre-Covid trust on the pandemic management. In other words, we expect a negative and statistically significant sign for our parameter of interest.

### 4.2 Identification strategy

One analysis that explores the effect of trust on the Covid-19 crisis may suffer from an endogeneity problem, namely, reverse causality. Indeed, the quality of crisis management can affect citizens’ trust in their government. One example is France, where statistics show substantial variations in citizens’ trust in their government since the start of the crisis due to health policies implemented. ^14^ More formal studies, such as Jennings (2020) and Schraff (2020), also point to the possible endogenous character of trust to the Covid-19 crisis. However, our analysis seems to be free of this endogeneity problem for two reasons: *i-*we consider, as previously stated, the pre-Covid trust; *ii-*Covid-19 is similar to a natural experiment that hit the world mainly in 2020, and it seems unlikely that the Covid crisis of the 2020-2021 period could influence the state of trust between 2018 and 2019 given the context-dependent character of responses to trust questions. Therefore, we can present the trust used in this work as exogenous to the Covid-19 crisis. Putting these two arguments together in addition to control variables used to address potential omitted relevant variable biases, we can consider our model estimated by Ordinary Least Squares (OLS) able to capture the causal effect of trust.

## 5 Results

Our baseline results are presented in Table 2. The first column presents the effect of trust on Covid-19 cases per million population. The results reveal a negative and significant effect of trust on the number of Covid-19 cases. More precisely, a one percent increase in trust decreases the number of infections per million population by 327.15. Relative to the sample mean, this result shows that a one percent increase in trust decreases the number of cases per million population by almost 2%. Finally, it is important to note that all our statistically significant control variables have the expected sign.

**Table 2.** Covid-19 crisis management and trust

| Table 2 – Covid-19 crisis management and trust |  |  |
| --- | --- | --- |
|  | [1]<br>Covid cases | [2]<br>Covid deaths |
| Trust in government | -327.148**<br>(136.3815) | -14.166***<br>(3.3913) |
| Population density | 19.269<br>(12.2138) | 0.558*<br>(0.3104) |
| Public expenditure | 156.269<br>(162.0462) | 3.839<br>(4.4129) |
| Hospital beds per thousand | -1402.661**<br>(571.7117) | -38.434**<br>(14.6180) |
| Trade openness | 145.250***<br>(32.5793) | 2.973***<br>(0.9046) |
| GDP per capita growth | -1345.276<br>(1421.9632) | -85.550**<br>(35.5362) |
| Observations | 41 | 41 |
| R-squared | 0.402 | 0.496 |

Unreported constant included. Robust standard errors in brackets. \*\*\* p<0.01, \*\* p<0.05, \* p<0.1.

In column [2], we analyze the effect of trust on Covid-19 deaths per million population. The result shows a negative effect of trust on the Covid-19 related deaths. The magnitude of the coefficient reveals that a one percent increase in trust decreases the number of deaths per million population by 14.16. Relative to the sample mean, this result indicates a decrease of almost 3% for a one percent increase in trust. As previously reported, all our control variables have the expected sign. ^15^

In sum, results in this section show that trust determines sound management of the pandemic. More concretely, on average, a country can experience a decrease in the number of cases and deaths per million population of 2% and 3% respectively for a one percent increase in the trust of citizens in their government.

## 6 Robustness

In this section, we mainly test the robustness of our results to alternative samples, additional controls, alternative definitions of the dependent and interest variables.

### 6.1 Alternative sample

We start our robustness exercise by testing the sensitivity of our results to sample selection. Four modifications of the sample are performed. First, we exclude March 1, 2020 to May 1, 2020 in computing the average number of cases and deaths. Indeed, this period is marked in many countries by stringent measures such as strict lockdown, which may overestimate the effect of trust on the pandemic management. Second, we take into account outliers by excluding the top (bottom) 5% of countries with high (low) trust in their government. ^16^ Finally, in order to reduce the heterogeneity ^17^ that may characterize the countries in our sample, we exclude the non-OECD countries to have a more homogeneous sample at least in terms of income levels. Results reported in columns [1]-[4] of Table 3 for the number of cases and deaths produce similar results to our baseline findings. In other words, our results are neither driven by strict lockdown measures nor outliers and income disparities.

**Table 3.**
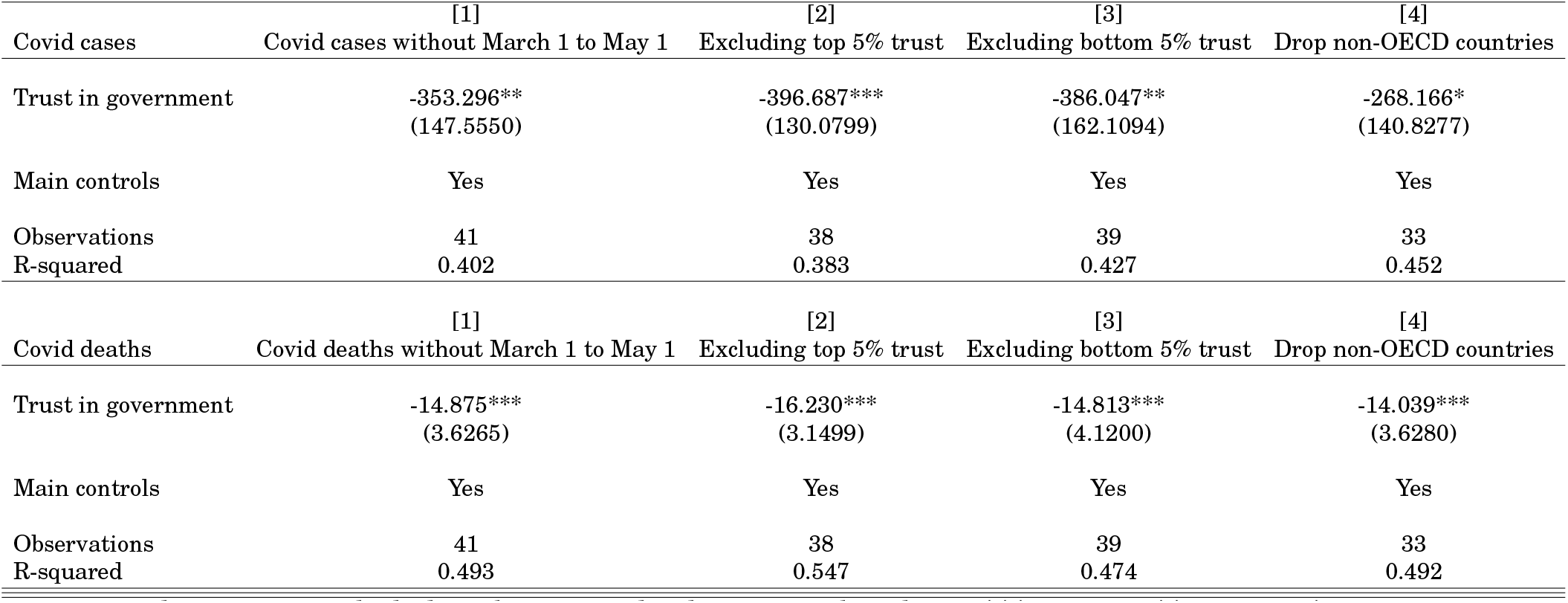
Alternative sample Unreported constant included. Robust standard errors in brackets. *** p<0.01, ** p<0.05, * p<0.1. Main controls are those in Table 2.

|  | [1] | [2] | [3] | [4] |
| --- | --- | --- | --- | --- |
| Covid cases | Covid cases without March 1 to May 1 | Excluding top 5% trust | Excluding bottom 5% trust | Drop non-OECD countries |
| Trust in government | -353.296**<br>(147.5550) | -396.687***<br>(130.0799) | -386.047**<br>(162.1094) | -268.166*<br>(140.8277) |
| Main controls | Yes | Yes | Yes | Yes |
| Observations | 41 | 38 | 39 | 33 |
| R-squared | 0.402 | 0.383 | 0.427 | 0.452 |

|  | [1] | [2] | [3] | [4] |
| --- | --- | --- | --- | --- |
| Covid deaths | Covid deaths without March 1 to May 1 | Excluding top 5% trust | Excluding bottom 5% trust | Drop non-OECD countries |
| Trust in government | -14.875***<br>(3.6265) | -16.230***<br>(3.1499) | -14.813***<br>(4.1200) | -14.039***<br>(3.6280) |
| Main controls | Yes | Yes | Yes | Yes |
| Observations | 41 | 38 | 39 | 33 |
| R-squared | 0.493 | 0.547 | 0.474 | 0.492 |

### 6.2 Potential omitted variables

We continue our robustness exercise by testing the sensitivity of our results to additional control variables. Based on the literature of the Covid-19 pandemic and trust, we include four groups of control variables. The first group includes a range of health policy variables and risk factors, including test policy (share of the population tested), positive rate, reproduction rate, share of the population vaccinated (vaccinations), the proportion of population above 65 years (above 65 yrs), the cardiovascular death rate (cardio. death), diabetes prevalence (diabetes), the share of the smoking population (smoke), and stringency index. The second group includes institutional variables such as democracy, government fractionalization (gov. frac.), years left in current term (yrs left in cur. term), government polarization (gov. polarization), and central bank independence (central bank ind.). The third group includes real economy and demographic variables such as inflation, unemployment, urbanization, level of development (level dev.), debt ratings (ratings), inequality, financial openness, and human capital. The fourth group includes social, migration, and health variables such as cultural tightness–looseness (tightness), the share of people adhering to religion (religion), ^18^ national pride, migrant stock (percent migrants), and pre-pandemic all-cause mortality (death). Detailed descriptions of these variables can be found in the Appendix. Results reported in columns [1]-[27] of Tables 4 and 5 show that including these variables yields similar results to our initial findings.

**Table 4.**
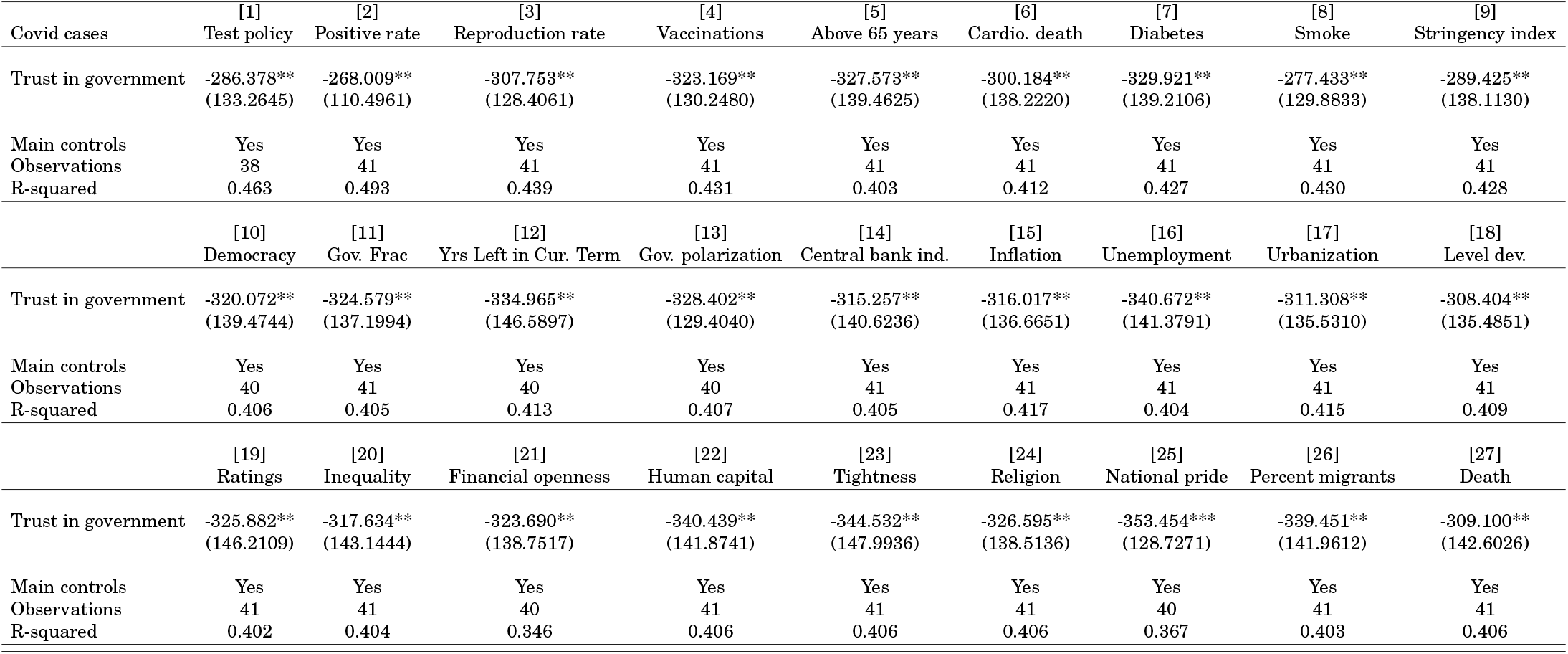
Potential omitted variables Unreported constant included. Robust standard errors in brackets. *** p<0.01, ** p<0.05, * p<0.1. Main controls are those in Table 2.

**Table 5.** Potential omitted variables Unreported constant included. Robust standard errors in brackets. *** p<0.01, ** p<0.05, * p<0.1. Main controls are those in Table 2.

|  | [1] | [2] | [3] | [4] | [5] | [6] | [7] | [8] | [9] |
| --- | --- | --- | --- | --- | --- | --- | --- | --- | --- |
| Covid deaths | Test policy | Positive rate | Reproduction rate | Vaccinations | Above 65 years | Cardio. death | Diabetes | Smoke | Stringency index |
| Trust in government | -13.817***<br>(3.3049) | -12.445***<br>(2.8427) | -13.065***<br>(2.5418) | -14.068***<br>(3.1250) | -14.162***<br>(3.4543) | -13.686***<br>(3.4908) | -14.225***<br>(3.5304) | -13.482***<br>(3.2730) | -13.299***<br>(3.4264) |
| Main controls | Yes | Yes | Yes | Yes | Yes | Yes | Yes | Yes | Yes |
| Observations | 38 | 41 | 41 | 41 | 41 | 41 | 41 | 41 | 41 |
| R-squared | 0.521 | 0.610 | 0.668 | 0.522 | 0.496 | 0.501 | 0.513 | 0.504 | 0.517 |
|  | [10] | [11] | [12] | [13] | [14] | [15] | [16] | [17] | [18] |
|  | Democracy | Gov. Frac | Yrs Left in Cur. Term | Gov. polarization | Central bank ind. | Inflation | Unemployment | Urbanization | Level dev. |
| Trust in government | -13.491***<br>(3.4644) | -14.372***<br>(3.8059) | -13.831***<br>(3.5120) | -13.697***<br>(3.2508) | -13.638***<br>(3.4933) | -14.024***<br>(3.4470) | -14.119***<br>(3.3381) | -13.703***<br>(3.5375) | -13.325***<br>(3.5215) |
| Main controls | Yes | Yes | Yes | Yes | Yes | Yes | Yes | Yes | Yes |
| Observations | 40 | 41 | 40 | 40 | 41 | 41 | 41 | 41 | 41 |
| R-squared | 0.508 | 0.526 | 0.502 | 0.496 | 0.503 | 0.500 | 0.496 | 0.512 | 0.517 |
|  | [19] | [20] | [21] | [22] | [23] | [24] | [25] | [26] | [27] |
|  | Ratings | Inequality | Financial openness | Human capital | Tightness | Religion | National pride | Percent migrants | Death |
| Trust in government | -15.474***<br>(3.3374) | -13.848***<br>(3.3517) | -14.223***<br>(3.2295) | -13.820***<br>(3.3645) | -14.221***<br>(3.5113) | -14.163***<br>(3.4429) | -14.808***<br>(2.8443) | -12.804***<br>(3.7413) | -13.088***<br>(3.5033) |
| Main controls | Yes | Yes | Yes | Yes | Yes | Yes | Yes | Yes | Yes |
| Observations | 41 | 41 | 40 | 41 | 41 | 41 | 40 | 41 | 41 |
| R-squared | 0.508 | 0.498 | 0.517 | 0.500 | 0.496 | 0.496 | 0.534 | 0.513 | 0.513 |

### 6.3 Alternative definitions of the dependent variable

In this section, we take a closer look at the definition of our dependent variables by changing the number of cases and deaths per million population (disease prevalence) in our model to new cases per million population (disease incidence), new deaths per million population (disease incidence), hospitalized patients per million population (hosp. patients), Intensive Care Unit (ICU) patients per million population, excess mortality (P-scores), and Covid Performance Index (CPI). The results are compiled in Table 6.

**Table 6.** Alternative definition of the dependent variable Unreported constant included. Robust standard errors in brackets. *** p<0.01, ** p<0.05, * p<0.1. Main controls are those in Table 2.

|  | [1]<br>New cases | [2]<br>New deaths | [3]<br>Hosp. patients | [4]<br>ICU patients | [5]<br>Excess mortality | [6]<br>CPI |
| --- | --- | --- | --- | --- | --- | --- |
| Trust in government | -2.293**<br>(0.9348) | -0.089***<br>(0.0188) | -4.205***<br>(1.3647) | -0.653***<br>(0.2011) | -0.381***<br>(0.0971) | 0.427*<br>(0.2396) |
| Main controls | Yes | Yes | Yes | Yes | Yes | Yes |
| Observations | 41 | 41 | 26 | 20 | 39 | 37 |
| R-squared | 0.426 | 0.467 | 0.631 | 0.662 | 0.433 | 0.431 |

They reveal that a high level of trust decreases the number of new cases and deaths and the number of hospitalized patients (columns [1]-[3]). The number of ICU patients also decreases (column [4]). A closer look at the magnitude of the coefficient shows that one standard deviation increase in trust decreases the number of ICU patients by 14.92 percentage points, i.e., a reduction of the unconditional mean by 60.7%. In addition, excess mortality is also negatively associated with trust in government (column [5]). Specifically, one standard deviation increase in trust decreases the excess mortality during the Covid-19 crisis by 3.78 percentage points, representing a decrease in the unconditional mean excess mortality by 32%. Finally, results regarding the effect of trust on Covid-19 performance show a positive and significant effect (column [6]). The size of the coefficient means that one standard deviation increase in trust increases the performance of the Covid-19 by 18.28 percentage points or an increase in the unconditional mean by 39%. In light of these results, we can easily say that changing the crisis management measures does not alter our conclusions.

### 6.4 Alternative definition of interest variable

Finally, we test the robustness of our results to alternative definitions of the interest variable in two ways.

First, instead of using the 2018-2019 average, we choose the average over the period 2006 (the latest year in our database) to 2019, the average over 2006-2019 without the Global Financial Crisis (GFC) period to purge our measure of the potential effect of the financial crisis on trust. Finally, and in contrast to previous measures, we take 2019 trust observations instead of average over any period. The results of these tests presented in columns [1]-[6] of Table 7 show negative and significant effects of the three measures of trust on cases (deaths) per million population, with coefficients close to the baseline model.

**Table 7.** Alternative definition of interest variable Unreported constant included. Robust standard errors in brackets. *** p<0.01, ** p<0.05, * p<0.1. Main controls are those in Table 2.

|  | [1]<br>Covid cases | [2]<br>Covid deaths | [3]<br>Covid cases | [4]<br>Covid deaths | [5]<br>Covid cases | [6]<br>Covid deaths | [7]<br>Covid cases | [8]<br>Covid deaths | [9]<br>Covid cases | [10]<br>Covid deaths | [11]<br>Covid cases | [12]<br>Covid deaths |
| --- | --- | --- | --- | --- | --- | --- | --- | --- | --- | --- | --- | --- |
| Trust over 2006-2019 | -367.388**<br>(164.0060) | -13.537***<br>(3.7042) |  |  |  |  |  |  |  |  |  |  |
| Trust over 2006-2019 w/o GFC |  |  | -379.403**<br>(160.7335) | -14.387***<br>(3.6017) |  |  |  |  |  |  |  |  |
| Trust in 2019 |  |  |  |  | -367.388**<br>(164.0060) | -13.537***<br>(3.7042) |  |  |  |  |  |  |
| Trust in politicians |  |  |  |  |  |  | -6290.975***<br>(1608.3302) | -244.255***<br>(57.4652) |  |  |  |  |
| Satisfaction with the government |  |  |  |  |  |  |  |  | -4757.442**<br>(1985.0109) | -260.601***<br>(59.6838) |  |  |
| Democracy satisfaction |  |  |  |  |  |  |  |  |  |  | -4494.167**<br>(1606.1738) | -215.779***<br>(39.4724) |
| Main controls | Yes | Yes | Yes | Yes | Yes | Yes | Yes | Yes | Yes | Yes | Yes | Yes |
| Observations | 41 | 41 | 41 | 41 | 41 | 41 | 24 | 24 | 24 | 24 | 24 | 24 |
| R-squared | 0.399 | 0.395 | 0.411 | 0.434 | 0.399 | 0.395 | 0.493 | 0.647 | 0.335 | 0.566 | 0.391 | 0.624 |

Second, we use trust measures from the European Social Survey (ESS) database, which provides survey data on trust in Europe. The survey consists of asking individuals to select between 0 (no trust) and 10 (total trust). For our work, and given the high number of intermediate answers between 0 and 10, it seems impossible to compute the proportion of people who declare trust in institutions as in our baseline model. For this reason, we rely on simple mean values by country of the different responses of the interviewees in Wave 9. The results based on three measures of trust, namely trust in politicians, satisfaction with the national government, and satisfaction with the way democracy works in the country, are presented in Table 7 (columns [7]-[12]) and show signs consistent with the baseline model. Indeed, high levels of these three trust measures are associated with low levels of Covid-19 cases (deaths). However, we must note that the magnitudes of the coefficients are much larger, probably due to the geographical coverage of the database (only in Europe) and to the difference in the scale of the trust variables compared to that used in the baseline model.

## 7 Heterogeneity

Previous results indicate that high trust between population and government favors successful management of the crisis through lower cases and deaths. In this section, we test the sensitivity of these results to the contamination level (cases) and the time dimension.

### 7.1 The level of contamination

In this section, we evaluate the sensitivity of our results to the number of cases per million population. To do so, we distinguish the *early* phase of the pandemic marked by the number of cases below 100 per million population from the *late* phase characterized by a level of contamination above this threshold. Results presented in column [1] of Table 8 reveal no evidence of trust effect on Covid-19 spread below 100 cases per million population. Concerning the number of deaths (column [2]), we observe a slight decrease with trust. However, beyond 100 cases per million population, the level of trust strongly determines the reduction of contamination and deaths (columns [3]-[4]). This result may be explained by higher compliance to containment measures that characterize high-trust countries once reaching this threshold. ^19^

**Table 8.**
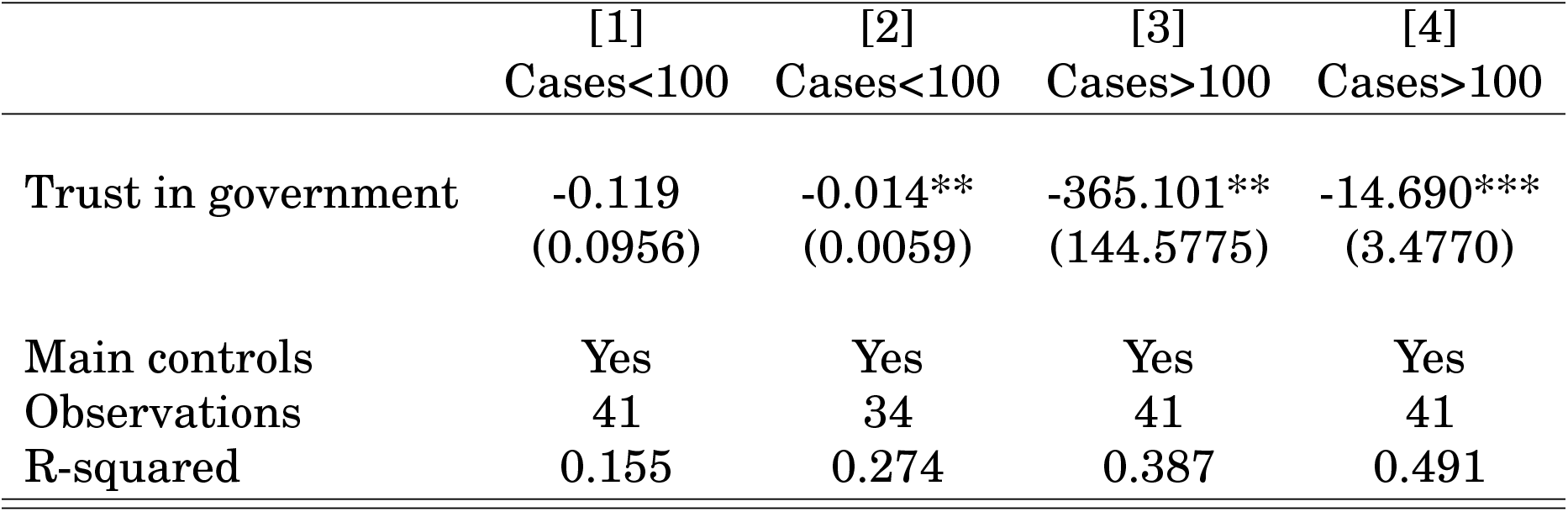
The level of contamination Unreported constant included. Robust standard errors in brackets. *** p<0.01, ** p<0.05, * p<0.1. Main controls are those in Table 2. In columns [1] and [3] ([2] and [4]), the number of cases (deaths) per million population is the dependent variable.

### 7.2 Time perspective

Attitudes of trust and other cultural and institutional traits may surprisingly persist for a long time (Acemoglu et al., 2001; Algan and Cahuc, 2007; Bjørnskov, 2007; Dear-mon and Grier, 2009; Tabellini, 2010). Capitalizing on this observation, this section investigates whether the effect of pre-crisis trust is long-lasting or short-lived. In other words, we document whether the effect of trust on crisis management ends in 2020 or keeps going. To do so, we analyze a dynamic effect of trust by assessing its impact on crisis management (number of cases and deaths per million population) in 2020 and 2021. The results presented in Table 9 show that trust influences the number of cases and deaths in both 2020 and 2021. More interestingly, the effect appears to increase over time. In other words, the effect in 2021, although our sample ends on May 26, 2021, already exceeds that found in 2020. In view of these results, it appears consistent with the literature that the effect of trust is more persistent than transitory in dealing with the crisis.

**Table 9.**
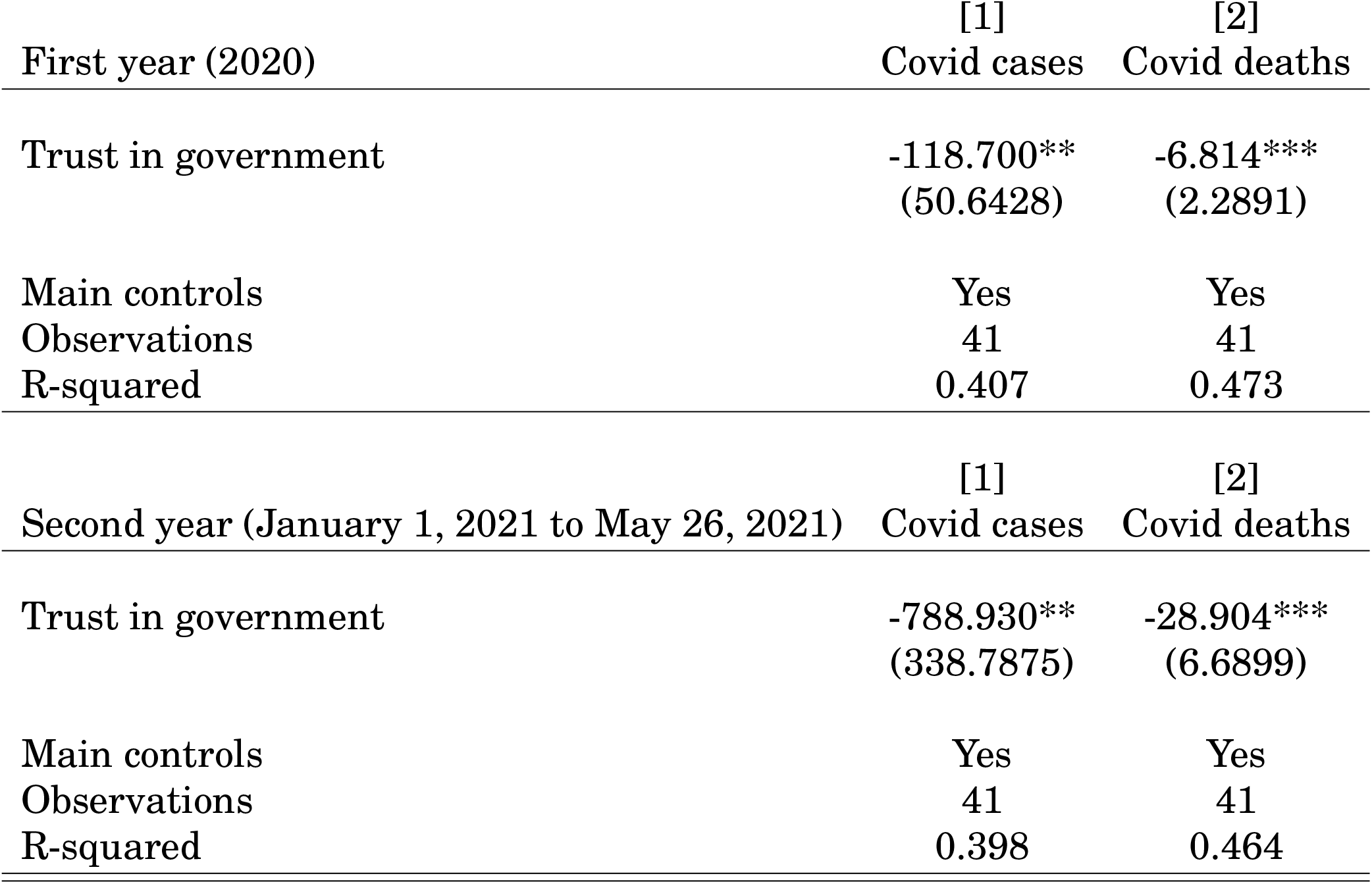
Time perspective Unreported constant included. Robust standard errors in brackets. *** p<0.01, ** p<0.05, * p<0.1. Main controls are those in Table 2.

## 8 Conclusion

In times when citizens’ commitment and responsibility are highly needed, this article documents the effect of pre-Covid trust in government on the pandemic’s management. Robust results from various tests, including alternative definitions of the dependent and interest variables, sample alteration, and additional controls, reveal that a high level of trust improves the Covid-19 crisis management through lower numbers of cases and deaths per million population. Heterogeneity tests performed later show that our results are sensitive to the level of contamination and increase over time. In the light of these different results, this paper calls for further work on trust between governments and citizens to increase compliance with the non-pharmaceutical and pharmaceutical health policies introduced to control the pandemic.

Amid various recovery plans, including the European plans of more than US$700 billion, the joint US$50 billion proposals from International Institutions such as the International Monetary Fund (IMF), the World Health Organization (WHO), the International Trade Organization (WTO) and the World Bank and the Paris summit announcements of Tuesday, May 18, this paper calls to take structural differences such as trust into account to achieve more effective management and more synchronized performance for less scattered recovery. In addition to the pandemic crisis, this paper could have important implications for other (current) crises, such as the climate and inequality crisis. Consequently, governments should invest more in trust, which has fallen especially in recent years, in order to increase tax compliance to support redistributive policies and achieve the Sustainable Development Goals (SDGs) on the one hand, and motivate citizens to fully comply with environmental policies on the other hand.

## Data Availability

Data are available on request

## Appendix

**Table 10.** Descriptive statistics of main variables

| Variable | Obs | Mean | Std. Dev. | Min | Max |
| --- | --- | --- | --- | --- | --- |
| Total cases per million | 41 | 20543.71 | 11637.1 | 377.94 | 47874.47 |
| Total deaths per million | 41 | 475.083 | 303.025 | 4.758 | 1211.676 |
| New cases per million | 41 | 137.865 | 79.056 | 1.222 | 343.45 |
| New deaths per million | 41 | 2.819 | 1.774 | 0.013 | 7.007 |
| ICU patients per million | 20 | 26.93 | 13.589 | 4.926 | 50.683 |
| Hosp. patients per million | 26 | 172.245 | 110.269 | 19.837 | 451.521 |
| Covid excess mortality | 39 | 12.204 | 10.871 | -3.057 | 55.672 |
| Covid Performance Index | 37 | 47.262 | 19.876 | 6.3 | 93 |
| Trust in government | 41 | 45.167 | 14.555 | 21.428 | 82.83 |
| Trust in politicians | 24 | 5.554 | 0.992 | 4.266 | 7.536 |
| Satisfaction with the government | 24 | 3.897 | 0.921 | 2.552 | 5.385 |
| Democracy satisfaction | 24 | 4.651 | 0.806 | 3.24 | 6.664 |
| Population density | 41 | 120.125 | 125.817 | 2.667 | 493.242 |
| Public expenditure | 41 | 40.227 | 9.497 | 17.35 | 54.386 |
| Hospital beds per thousand | 41 | 4.445 | 2.647 | 1.13 | 13.05 |
| Trade openness | 41 | 82.764 | 47.941 | 23.194 | 285.96 |
| GDP per capita growth | 41 | 2.076 | 1.308 | 0.561 | 5.481 |

**Table 11.** List of countries

| Table 11 – List of countries |  |
| --- | --- |
| Country | Country |
| Australia | Latvia |
| Austria | Lithuania |
| Belgium | Luxembourg |
| Brazil | Mexico |
| Canada | Netherlands |
| Chile | New Zealand |
| Colombia | Norway |
| Costa Rica | Poland |
| Czechia | Portugal |
| Denmark | Russia |
| Estonia | Slovakia |
| Finland | Slovenia |
| France | South Africa |
| Germany | South Korea |
| Greece | Spain |
| Hungary | Sweden |
| Iceland | Switzerland |
| Ireland | Turkey |
| Israel | United Kingdom |
| Italy | United States |
| Japan |  |

## Footnotes

1. See https://www.bbc.com/news/world-52103747 for various albeit similar policies implemented by countries.

2. For example, and as illustrated by Table 10 in the Appendix, the number of cases and deaths per million population varies widely, with differences between the maximum and minimum of 47496.53 and 1206.92 respectively.

3. Indeed, trust is identified in the literature as an essential precondition for successful regulation and people’s ability to comply with the rule of law (see for instance https://www.oecd.org/gov/trust-in-government.htm. In current times where regulation of the economy and compliance to government guidelines appears necessary, trust appears to be very important

4. In this paper, trust in government and trust are used interchangeably.

5. While it is reasonable to think that cultural tightness-looseness and trust in government may be “strongly” related (see for instance Aktas et al., 2016; Su et al., 2016 or https://harvardpolitics.com/culture-response-covid-19/), statistical tests seem to tell a fairly opposite story with a nonsignificant correlation coefficient of 0.20 for countries in our sample. Later in the paper, we control for cultural tightness-looseness to better isolate the effect of trust in government on the pandemic management.

6. Furthermore, corruption, i.e., poor institutional quality, is another serious source of distrust between governments and the governed. Thus, higher trust between governments and citizens can arise from the better institutional quality, particularly through better corruption levels, paving the way for a prepandemic institutional situation suitable for transparent and efficient pandemic management.

7. https://knowablemagazine.org/article/society/2021/danger-high-public-debt-is-not-what-you-think

8. For example, in France, in order to limit these damages, some devices such as specialists’ consultation like psychologists were put in place.

9. See https://wellbeingtrust.org/areas-of-focus/policy-and-advocacy/reports/projected-deaths-of-despair-during-covid-19/ which also exposes collateral damage and amplifying factors of the crisis, in particular the “deaths of despair” tragedy caused by the implementation of various containment policies. Note that three factors, already at work, are exacerbating “deaths of despair”: unprecedented economic failure paired with massive unemployment mandated social isolation for months and possible residual isolation for years, and uncertainty caused by the sudden emergence of a novel, previously unknown microbe.

10. In other words, we use a pre-crisis measure of trust that remains completely unaffected by Covid-19 crisis management. This choice allows us to capture the role of differences in civic norms and trust in the political system that existed in the countries in our sample.

11. See also https://ourworldindata.org/covid-excess-mortality and https://www.who.int/data/stories/the-true-death-toll-of-covid-19-estimating-global-excess-mortality

12. Note that trust and level of development are strongly related. To take this into account and produce more informative statistics, we use a residual approach by adjusting trust to the level of development.

13. The classification of countries is based on the position in relation to the sample median. Thus, countries with a higher trust level are those above the sample median, and those below the median are classified as having a lower trust level.

14. See :http://www.statista.com/statistics/1107643/covid-19-trust-government-france/

15. Based on conclusions of Table 1 on the one hand and, on the other hand, for a better appreciation of these results, we evaluate the effect according to the level of trust. In other words, we compare countries with high trust to countries with low trust. To do so, we compute a dummy variable that takes 1 (high trust countries) if country *i*’s trust is above the sample median and 0 (low trust countries) for countries whose trust is below the sample median and re-estimate our baseline model by replacing trust with this new (dummy) variable. The results, available upon request, show that high-trust countries exhibit lower numbers of cases and deaths per million population (8143.90 and 358.38 respectively) compared to lowtrust countries. Applied to France, which has a population of 67.06 million based on 2020 estimations (see https://www.insee.fr/fr/statistiques/4277615?sommaire=4318291), this would amount to a reduction in the number of cases and deaths of 546129.93 and 24032.96 respectively.

16. The top 5% high trust countries are Luxembourg, New Zealand, Switzerland and the bottom 5% low trust countries are Italy and Latvia

17. Another way to reduce cross-country heterogeneity (and subsequently have a relatively normal distribution of variables) is to use a log-transformation for the crisis management variables, notably the number of cases and deaths. Results of this transformation, available on request, show a negative and significant effect of trust in government on the logarithm of the number of cases and deaths. Specifically, a one percent increase in citizens’ trust in their government reduces the number of cases and deaths by 3% and 5% respectively.

18. Religion affiliation may play a significant role in trust and values, as pointed out by Guiso et al., 2006 and Ortiz-Ospina and Roser, 2016.

19. It is important to note that below 100 cases per million population, many countries do not implement effective containment measures, which may justify virtually the same pattern of crisis management in this phase of the pandemic, i.e., statistically non-significant effect of trust on the number of cases and a relatively small effect on the number of deaths. In addition, existing literature, including Fotiou and Lagerborg (2021) shows high efficiency of health or containment policies once 100 cases per million of population are reached.

